# Characteristics and outcomes of clinically diagnosed RT-PCR swab negative COVID-19: a retrospective cohort study

**DOI:** 10.1101/2020.10.20.20204651

**Authors:** Paul Middleton, Pablo N. Perez-Guzman, Alexandra Cheng, Naveenta Kumar, Mara D. Kont, Anna Daunt, Sujit Mukherjee, Graham Cooke, Timothy B. Hallett, Katharina Hauck, Peter J. White, Mark R. Thursz, Shevanthi Nayagam

**Author notes:** Corresponding Author: Dr Shevanthi Nayagam, MRC Centre for Global Infectious Disease Analysis Department of Infectious Disease Epidemiology, Faculty of Medicine at St Mary’s Campus, Imperial College London, W2 1PG. UK. These first authors contributed equally to this article (Paul Middleton, Pablo N. Perez-Guzman, Shevanthi Nayagam). The opinions expressed by authors contributing to this journal do not necessarily reflect the opinions of the Centers for Disease Control and Prevention or the institutions with which the authors are affiliated. Conflicts of interest: None.

## Abstract

Patients with strong clinical features of COVID-19 with negative real time polymerase chain reaction (RT-PCR) SARS-CoV-2 testing are not currently included in official statistics. The scale, characteristics and clinical relevance of this group are thus unknown. We performed a retrospective cohort study in two large London hospitals to characterize the demographic, clinical, and hospitalization outcome characteristics of swab-negative clinical COVID-19 patients. We found 1 in 5 patients with a negative swab and clinical suspicion of COVID-19 received a clinical diagnosis of COVID-19 within clinical documentation, discharge summary or death certificate. We compared this group to a similar swab positive cohort and found similar demographic composition, symptomology and laboratory findings. Swab-negative clinical COVID-19 patients had better outcomes, with shorter length of hospital stay, reduced need for >60% supplementary oxygen and reduced mortality. Patients with strong clinical features of COVID-19 that are swab-negative are a common clinical challenge. Health systems must recognize and plan for the management of swab-negative patients in their COVID-19 clinical management, infection control policies and epidemiological assessments.

## Background

As of 2^nd^ August 2020, the World Health Organization (WHO) has reported 17.6 million confirmed cases of COVID-19 globally with 532,340 confirmed deaths.[1] WHO defines a confirmed case as a person with laboratory confirmation of COVID-19 infection.[2] Cases where laboratory confirmation is not done or inconclusive are recognized as probable cases however those with strong clinical features, but negative testing are not recognized.[2] Epidemiological evaluations in many countries, including the UK, focus on patients with positive real-time polymerase chain reaction (RT-PCR) testing.[3] This places a significant importance on the diagnostic accuracy of laboratory testing when it has been performed.

However, the diagnostic accuracy of RT-PCR upper respiratory tract swabs is increasingly being questioned. A study utilizing both chest computerized tomography imaging (CT) and RT-PCR testing in patients with suspected COVID-19 found 75% of cases with a negative RT-PCR test had CT findings suggestive of COVID-19.[4] Several case reports describe patients with clinical features of COVID-19 but negative upper respiratory tract swabs who later have positive confirmatory testing on induced sputum or bronchial lavage.[5–7] A recent analysis of published cohorts calculated the false-negative rate of RT-PCR testing amongst symptomatic COVID-19 patients who eventually test positive for COVID-19 is likely 20% and increases gradually with time since symptoms onset.[8]

Acknowledging this, some patients with strong features of COVID-19 receive a clinical diagnosis of COVID-19 despite a negative swab result. This clinical approach is being further recognized in the admission criteria of some clinical trials who permit recruitment of these patients.[9] For example in the recently reported RECOVERY trial 10% of those randomized to dexamethasone had a negative swab at the time of randomization.[10] However, little has been documented about the scale and clinical relevance of this subgroup of patients for whom there is no consensus on optimal management. Our study aims to assess the real-world prevalence and characteristics of clinically diagnosed swab-negative COVID-19, including factors associated with swab-negative disease, and whether their outcomes differ to swab-positive patients.

## Methods

We retrospectively reviewed medical admissions from March 1 to April 12, 2020 at Imperial College Healthcare Trust (ICHNT) in London, UK from two admitting sites. We defined eligible cases as those who presented with clinical suspicion of COVID-19 or had symptoms compatible with COVID-19, were admitted to hospital and had a SARS-CoV-2 nasopharyngeal swab performed. We collected full demographic characteristics, time course of symptoms, time of presentation and testing, presenting symptoms, final diagnosis and outcome as well as radiological and laboratory findings for all patients with a negative swab from admission until discharge. Cases were evaluated using the Public Health England (PHE) testing criteria for possible COVID cases.

We defined swab-negative clinical COVID-19 cases as follows: a) clinical COVID-19 or high level of suspicion as defined by the treating medical team (as recorded in the medical notes, discharge documents or death certificate); and b) RT-PCR swab-negative (on initial and any subsequent tests performed).

The swab-negative clinical COVID-19 group of patients was compared to a subgroup of a previously described swab-positive cohort who were similarly admitted via general medical admissions.^11^ Chi-square, Fisher’s and rank sum tests were used, as appropriate, to compare the cohorts’ characteristics, and odds ratios (OR) calculated to assess differences in the outcomes of respiratory deterioration, defined as requiring greater than 60% oxygen, and death. Lastly, we assessed the cumulative risk of the competing outcomes of hospital discharge and death over time using the using the Nelson-Aelen estimator.

The study was approved by the ICHNT clinical governance team. As we report on routinely collected non-identifiable clinical audit data, no individual informed consent was required under the UK policy framework for health and social care. All methods were conducted in accordance with relevant guidelines and regulations.

## Results

We identified 1,119 emergency medical admissions with initial clinical suspicion of COVID-19 who had a SARS-CoV-2 swab performed (Figure 1). Initial swab was negative in 456 (41%) and positive in 663 (59%) patients. 62% (281/456) of those who were swab-negative received an alternative diagnosis. 47 of the swab-negative cohort later tested positive for COVID-19 on repeat PCR testing. The most common alternative diagnoses included community acquired pneumonia, lower respiratory tract infection and exacerbations of chronic lung disease such as chronic obstructive pulmonary disease.

**Figure 1.**
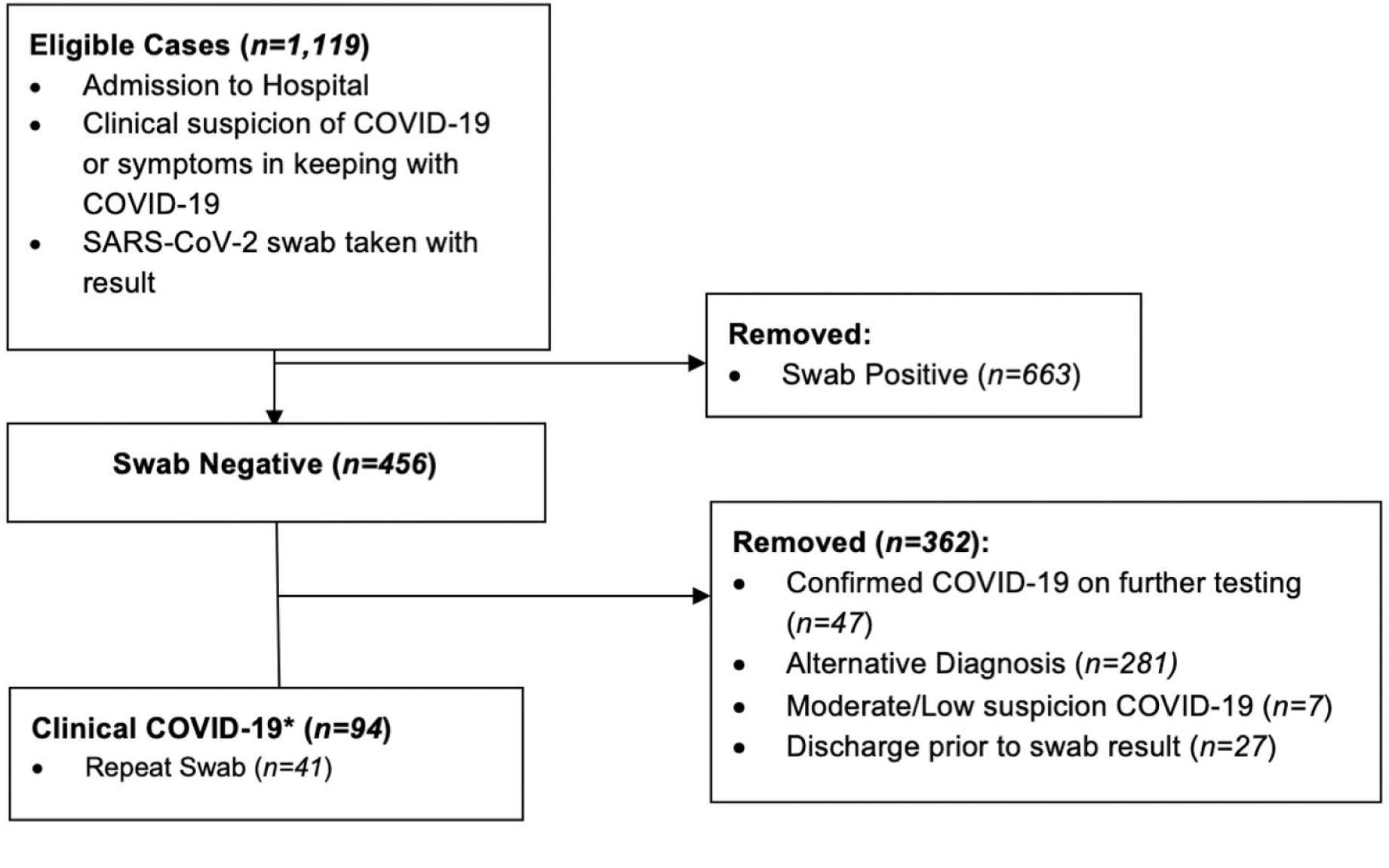
Case identification. *98% (92/94) Clinical COVID-19 cases fulfilled PHE guidance on testing eligibility

Overall, 20% (94/456) of swab-negative cases were identified as swab-negative clinical COVID-19. 98% (92/94) of these cases fulfilled PHE criteria for patients eligible to be swabbed for COVID-19 compared to 84% of the entire swab-negative group. 41/94 had repeat swab testing performed and remained negative.

The demographic profile and symptomatology of the swab-positive and swab-negative cohorts were similar, with high rates of influenza-like symptoms, cough and fever in both groups (Table). Typical features such as shortness of breath was higher in the swab-negative clinical COVID-19 cohort (75/92 (79.79%) vs 312/478 (66.67%) p=0.017). The swab-negative cohort was more likely to have chest radiographs reporting typical COVID-19 appearances than the swab-positive cohort (53/77 (68.83%) vs 195/389 (50.13%); p=0.004).

**Table 1.** Description of clinical characteristics and clinical course. *Abbreviations: CK, creatine kinase; CRP, C-reactive protein; EWS, early warning score; FiO2, inspiratory fraction of oxygen; IQR, interquartile range; LDH, lactate dehydrogenase*. * Pearson’s Chi-squared test with Yates’ continuity correction ^†^ Wilcoxon rank sum test with continuity correction ^‡^ Fisher’s exact test for count data

|  | RT-PCR-positive<br>(n = 468) | RT-PCR-negative<br>(n = 94) | p value |
| --- | --- | --- | --- |
| **Demography** |  |  |  |
| Male, n (%) | 288 (61.5%) | 57 (60.6%) | 0.962 * |
| Median age (IQR) | 68 (54 - 79) | 67 (54.25 - 78) | 0.638 † |
| **Symptoms** |  |  |  |
| Influenza-like syndrome, n (%) | 341 (72.9%) | 74 (78.7%) | 0.293 * |
| Cough, n (%) | 353 (75.4%) | 71 (75.5%) | 1.000 * |
| Fever, n (%) | 389 (83.1%) | 75 (79.8%) | 0.530 * |
| Nasal discharge, n (%) | 9 (1.9%) | 3 (3.2%) | 0.433 ‡ |
| Shortness of breath, n (%) | 312 (66.7%) | 75 (79.8%) | **0.017** * |
| Wheezing, n (%) | 1 (0.2%) | 4 (4.3%) | **0.003** ‡ |
| Anosmia, n (%) | 12 (2.6%) | 10 (10.6%) | **0.001** * |
| **Admission Oxygen Requirements (FiO2)** |  |  |  |
| Room air, n (%) | 254 (54.3%) | 50 (53.2%) | 0.937 * |
| 24 to <40% | 108 (23.1%) | 31 (33.0%) | 0.058 * |
| 40 to <60% | 8 (1.7%) | 3 (3.2%) | 0.405 ‡ |
| >60% | 98 (20.9%) | 10 (10.6%) | **0.030** * |
| **Chest Radiograph** |  |  |  |
| Normal CXR | 65/389 (16.7%) | 7/77 (9.1%) | 0.129 * |
| Typical COVID-19 CXR | 195/389 (50.2%) | 53/77 (68.8%) | **0.004** * |
| Atypical COVID-19 CXR | 106/389 (27.3%) | 15/77 (19.5%) | 0.201 * |
| Non-COVID abnormal CXR | 23/389 (5.9%) | 2/77 (2.6%) | 0.403 ‡ |
| **Laboratory Test** |  |  |  |
| Lymphocytes ≥1.1 | 154/460 (33.5%) | 25 (26.6%) | 0.238 * |
| Lymphocytes 0.5-1.0 | 258/460 (56.1%) | 65 (69.2%) | **0.026** * |
| Lymphocytes <0.5 | 43/460 (9.4%) | 4 (4.3%) | 0.152 ‡ |
| CRP ≥100 | 230/448 (51.3%) | 55/93 (59.1%) | 0.209 * |
| CRP 10-99 | 186/448 (41.5%) | 33/93 (35.5%) | 0.336 * |
| CRP <10 | 32/448 (7.1%) | 5/93 (5.4%) | 0.698 * |
| D-dimer ≥3000 | 42/225 (18.7%) | 16/66 (24.2%) | 0.411 * |
| D-dimer 2000-2999 | 29/225 (12.9%) | 9/66 (13.6%) | 1.000 * |
| D-dimer 1000-1999 | 64/225 (28.4%) | 19/66 (28.8%) | 1.000 * |
| D-dimer 500-999 | 63/225 (28.0%) | 18/66 (27.3%) | 1.000 * |
| D-dimer <500 | 27/225 (12.0%) | 4/66 (6.1%) | 0.255 ‡ |
| LDH ≥243 | 166/179 (92.7%) | 48/51 (94.1%) | 0.976 * |
| LDH <243 | 13/179 (7.3%) | 3/51 (5.9%) | 1.000 ‡ |
| CK ≥320 | 63/210 (30.0%) | 16/64 (25.0%) | 0.538 * |
| CK <320 | 147/210 (70.0%) | 48/64 (75.0%) | 0.538 * |
| Ferritin ≥5000 | 13/259 (5.0%) | 4/68 (5.9%) | 0.761 ‡ |
| Ferritin 1000-4999 | 94/259 (36.3%) | 27/68 (39.7%) | 0.706 * |
| Ferritin 500-999 | 73/259 (28.2%) | 15/68 (22.1%) | 0.390 * |
| Ferritin 300-499 | 38/259 (14.7%) | 10/68 (14.7%) | 1.000 * |
| Ferritin <300 | 41/259 (15.8%) | 12/68 (17.7%) | 0.860 * |
| **Clinical Course** |  |  |  |
| Median (IQR) days prior to admission | 6 (3 - 10) | 7 (4.25 - 13) | **0.013** † |
| Median (IQR) length of stay | 6 (4 - 10) | 5 (2.25 - 6) | **<0.001** † |
| Received ≥60% FIO2, n (%) | 215 (45.9%) | 25 (26.6%) | **0.001** * |
| Died, n (%) | 151 (32.3%) | 15 (16.0%) | **0.002** * |

Overall haematological and biochemical findings that have been associated with COVID-19 including raised CRP, d-dimer and ferritin were similarly distributed between the two cohorts. A higher proportion of patients had moderate lymphopenia within the swab-negative group (65/94 (69.15%) vs 258/460 (56.09%); p=0.026).

Patients in the swab-negative group presented slightly later in their clinical course after symptom onset, at a median of 7 days (IQR 4.3-13) for the RT-PCR-negatives compared to 6 days (IQR 3-10) within the RT-PCR-positives cohort (p<0.001). Interestingly, RT-PCR-negative patients had better hospitalisation outcomes than RT-PCR-positives. The former had a 57% lower probability of requiring ≥60% supplementary oxygen during hospitalisation (OR 0.43, 95%CI 0.26-0.70, p<0.001), a shorter length of hospital stay (median 5 days vs 6 days, p<0.001) and 60% lower probability of death (OR 0.40, 95%CI 0.22-0.72, p<0.001). The cumulative risk of being discharged alive was significantly higher in the swab-negative group than the positive group (p < 0.001). Furthermore, the swab-negative cohort have a lower cumulative risk of in-hospital mortality than positives (p<0.001) (Figure 2).

**Figure 2.**
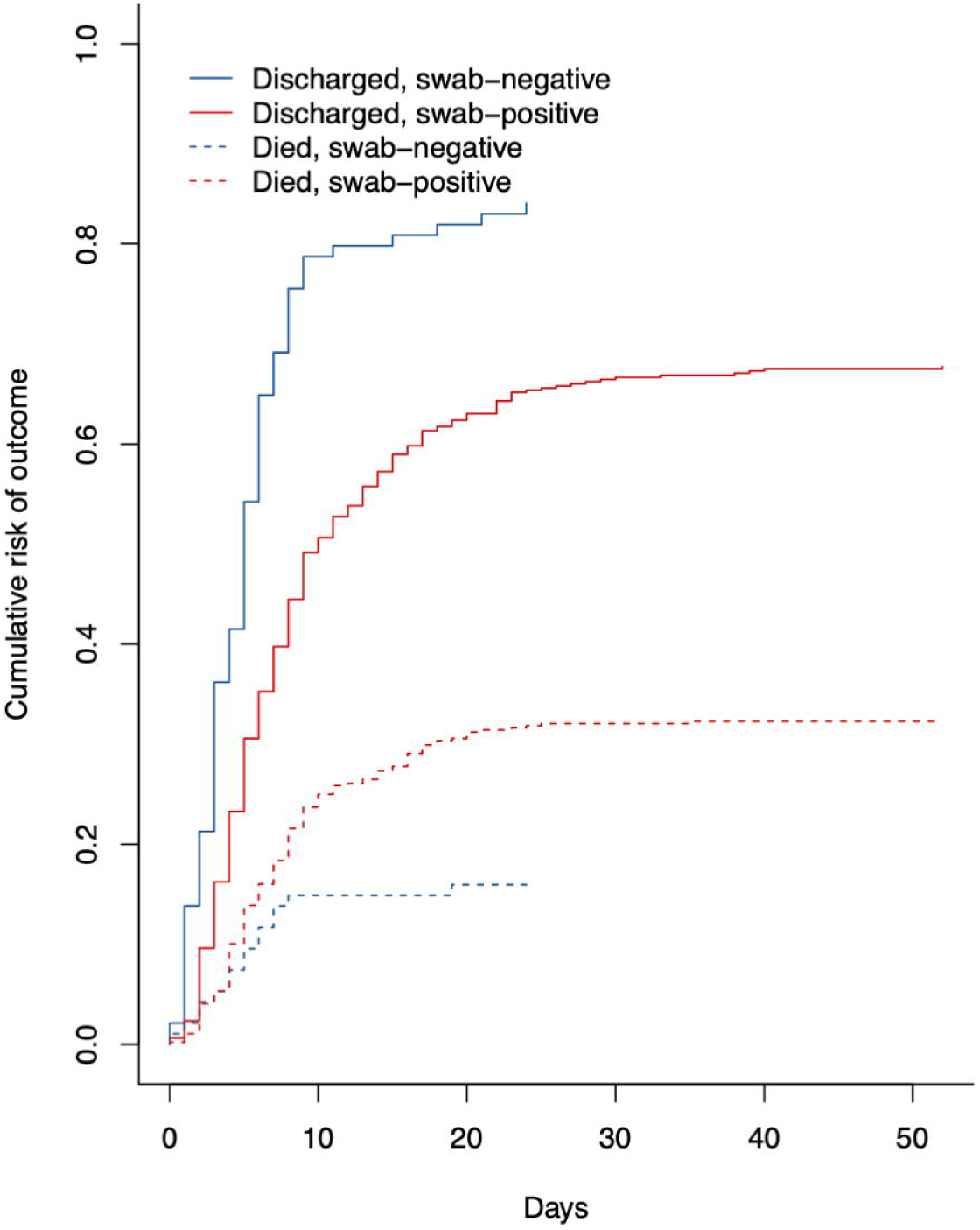
Cumulative risk of hospitalization outcomes by swab status.

## Discussion

We find one in five patients who had a negative COVID-19 swab received a clinical diagnosis of COVID-19 despite negative testing. This represents a 13% higher estimate of the number of hospitalized COVID-19 patients compared to considering swab-positive patients alone. Overall the clinical, biochemical and radiological features of the swab-negative clinical COVID-19 cohort did not differ significantly to the swab-positive cohort. Typical features of COVID-19 such as shortness of breath, typical chest x-ray findings and lymphopenia are more common in the swab-negative cohort as we would expect in a group defined by these parameters. Despite these similarities, swab-negative patients tended to have a longer delay between symptom onset and hospitalization, and had significantly better hospitalization outcomes, including mortality. Whether this potentially less aggressive phenotype is related to a lower viremia, variations in immune response or a different stage of illness presenting after period of peak viral replication is not yet known. Further research including immunological profiling would help further elucidate underlying disease mechanisms.

Our study is, to our knowledge, the first to detail a cohort of swab-negative clinical COVID-19. Our results provide evidence to validate swab-negative COVID-19 as a real clinical entity commonly encountered in hospital settings, findings which have important implications for current clinical practice and public health guidance.[12]

Firstly, it highlights that clinicians caring for COVID-19 patients should maintain a high clinical suspicion, even in the presence of a negative swab result. Our results suggest that a combination of clinical, radiological and biochemical features in keeping with COVID-19 disease, rather than swab results alone, should guide clinical management.

Secondly, managing swab-negative clinical COVID-19 within hospital poses disease control challenges. Cohorting this group with swab-positive patients presents potential risk of SARS-CoV-2 infection, if the clinical diagnosis is incorrect.^12^ Alternatively, isolating these patients from both swab-negative low-risk patients and swab-positive groups would place significant strain on hospital resources and may be unfeasible during an outbreak. Repeated testing may be beneficial in some cases however in our cohort 41/94 swab negative clinical COVID-19 cases had a repeat swab which remained negative. Testing on sputum or bronchial lavage may provide greater sensitivity.^5–7^ However, their use is limited by feasibility and potential risk to healthcare workers. Serological testing to confirm exposure to SARS-CoV-2 is likely the optimal second-line test as it is not operator-dependent, is low-risk to healthcare workers, and can feasibly be performed on a large number of patients. Serological proof of exposure could reduce concern regarding cohorting with swab-positive patients. However, these tests are not yet in routine clinical practice and require further evaluation.

Lastly, but importantly, from a public health perspective, swab-negative clinically diagnosed COVID-19 patients may not be captured in surveillance statistics, thereby underestimating healthcare demand. As the pandemic evolves countries with reducing incidence should consider this clinical group and how they should be addressed when planning interventions to prevent second-wave outbreaks such as contact tracing.

Our study is limited by its retrospective nature and likely under-estimates the proportion of patients with swab-negative clinical COVID-19, as it requires a high degree of clinical confidence without alternative diagnosis. The lack of positive confirmatory test means we cannot be sure of the true diagnosis in the clinically diagnosed swab negative group. However, this does not detract from the clinical problems this group presents to hospital management and epidemiological assessment. In the absence of improved testing or routine second line testing for patients who have high clinical suspicion of COVID-19 but negative swab testing, this group is likely to continue to pose a clinical challenge. During the period of data collection changes in local policy regarding repeated testing prevented re-testing for COVID-19 even in cases with high clinical suspicion therefore not all patients had repeated testing performed.

Patients with strong clinical features of COVID-19 who have negative nasopharyngeal RT-PCR test results are a common but understudied clinical group. Patients with a clinical diagnosis of COVID-19 do not differ significantly to similar swab-positive patients but seem to have better outcomes. Healthcare services should recognize and plan for the management of this group when making disease control interventions and epidemiological assessments.

## Data Availability

Individual patient data is not available

## Author Contributions

SN – Conceived study, data interpretation and provided clinical expertise and critical revision of the manuscript for intellectual and scientific content

PM – Conceived study, wrote the first manuscript and led on acquisition of data, interpreted data, provided clinical expertise and critical revision of the manuscript for intellectual and scientific content

PP – Conceived study, performed statistical analysis, interpreted data, provided clinical expertise and critical revision of the manuscript for intellectual and scientific content

AC – Supported data acquisition and curation and critical revision of the manuscript for intellectual and scientific content

NK - Supported data acquisition and curation and critical revision of the manuscript for intellectual and scientific content

AD - Supported data acquisition and curation and critical revision of the manuscript for intellectual and scientific content

SM - Supported data acquisition and curation and critical revision of the manuscript for intellectual and scientific content

MDK – Performed statistical analysis, data interpretation and critical revision of the manuscript for intellectual and scientific content

GC – Provided clinical expertise and critical revision of the manuscript for intellectual and scientific content

MRT – Provided clinical expertise and scientific expertise to lead the discussion of clinical and public health implications and critical revision of the manuscript for intellectual and scientific content

PJW - Provided clinical expertise and scientific expertise to lead the discussion of clinical and public health implications and critical revision of the manuscript for intellectual and scientific content

TBH - Provided clinical expertise and scientific expertise to lead the discussion of clinical and public health implications and critical revision of the manuscript for intellectual and scientific content

KH - Provided clinical expertise and scientific expertise to lead the discussion of clinical and public health implications and critical revision of the manuscript for intellectual and scientific content

## Acknowledgements

SN, TBH, KH, PJW and PP are supported by the MRC Centre for Global Infectious Disease Analysis (MR/R015600/1); this award is jointly funded by the UK Medical Research Council (MRC) and the UK Department for International Development (DFID) under the MRC/DFID Concordat agreement and is also part of the EDCTP2 program supported by the European Union (EU). The authors would also like to acknowledge the support of the Imperial BRC Centre and J-IDEA. The authors would like to thank all patients and staff at Imperial College Healthcare NHS Trust.

Role of the Funder/Sponsor: The funding organizations had no role in the design and conduct of the study; collection, management, analysis, and interpretation of the data; preparation, review, or approval of the manuscript; and decision to submit the manuscript for publication.

